# A novel plasma proteomic signature predicts long-term incident heart failure risk among persons living with HIV

**DOI:** 10.64898/2026.01.08.25343149

**Authors:** Saate S Shakil, Tristan Grogan, Kaku So-Armah, Peter Ganz, Matthew S Freiberg, Priscilla Y Hsue

**Affiliations:** Department of Medicine, University of California Los Angeles, CA, USA; Department of Medicine, Boston University, Boston, MA, USA; Department of Medicine, University of California San Francisco, CA, USA; Department of Medicine, Vanderbilt University, Nashville, TN, USA

**Author notes:** Corresponding author: SSS, Division of Cardiology, 1100 Glendon Ave Ste 1820, Los Angeles, CA 90401.

## Abstract

**Background:** Persons with HIV (PWH) have an increased risk of heart failure compared with the general population despite effective viral suppression. No methods currently exist to predict heart failure risk in PWH. We sought to develop a proteomics-based risk model of incident heart failure in a longitudinal cohort of PWH in the Veterans Aging Cohort Study Biomarker Cohort.

**Methods:** We characterized the baseline plasma proteome of 1,398 PWH using the SomaScan 4.0 aptamer-based platform. We incorporated traditional risk factors, HIV-specific variables, and inflammatory/coagulation markers into Cox proportional hazards models of incident heart failure. For protein risk models, we used least absolute shrinkage and selection operator (LASSO) regression with 10-fold cross-validation to identify the optimal set of predictors. Model performance was evaluated using Harrell’s Concordance Index (C-index).

**Results:** Among 1,398 PWH, 240 (17.1%) developed incident heart failure over 13.1 years of median follow-up. The sample was randomly divided 2:1 into training (n = 932) and test (n = 466) sets. In the training set, 4,248 proteins were screened, identifying 227 significant markers (Bonferroni-corrected p-value <1E-5); 108 of these were validated in the test set (Bonferroni-corrected p <2E-4) and included as candidate predictors in downstream LASSO models. Using LASSO-Cox regression, we identified a subset of predictor proteins alone and in combination with clinical and laboratory risk factors. Among the clinical risk factor models, individual traditional risk factors in combination with HIV disease markers (CD4^+^ T-cell count, viral load) exhibited the best predictive performance (C-index 0.656 [95% confidence interval 0.591 – 0.720]). Addition of circulating inflammatory and coagulation markers (soluble CD-14, interleukin-6, D-dimer) did not substantially improve performance (C-index 0.657 [0.594 – 0.720]). By contrast, a protein-only model incorporating 8 markers achieved an improved C-index of 0.725 [0.597 – 0.825]; proteins vs clinical model, p = 0.033), while the addition of CD4^+^ count and viral load performed similarly (C-index 0.715 [0.587 – 0.816]).

**Conclusion:** An 8-protein signature demonstrated superior predictive performance for long-term incident heart failure risk among PWH compared with baseline traditional risk factors and inflammatory markers. These findings highlight the potential of a proteomics-based approach for risk stratification in this population, which provides additive value beyond traditional or HIV-specific clinical risk factors.

## Introduction

Despite effective viral suppression with antiretroviral therapy, persons with HIV (PWH) suffer from an increased risk of heart failure compared with the general population, independently of the presence of coronary artery disease.^1,2^ While traditional cardiometabolic comorbidities contribute to this risk,^3–5^ HIV-specific immune dysregulation also predisposes to cardiovascular disease and mortality, with increased monocyte and T-cell activation and other markers of immune dysfunction, inflammation, and coagulation (e.g., sCD14, interleukin-6, D-dimer).^6–9^ These pathways are believed to lead to myocardial inflammation, steatosis, and fibrosis predisposing to adverse remodeling and both systolic and diastolic clinical heart failure.^2,10–14^ However, the exact mechanisms underlying heart failure in PWH remain largely uncharacterized.

PWH with heart failure experience higher all-cause mortality compared with those without HIV with heart failure and may therefore benefit from tailored screening and risk stratification.^15^ However, current heart failure risk prediction tools have largely been developed in the general population, including the Pooled Cohort Equations to Prevent Heart Failure (PCP-HF) and the American Heart Association Predicting Risk of CVD EVENTs (PREVENT) equations.^16,17^ Further, multiple studies have demonstrated that cardiovascular risk prediction algorithms derived in the general population perform poorly in PWH.^18–20^ Currently, there are no available risk calculators to stratify PWH at highest risk of developing heart failure. Precision approaches that account for unique HIV-specific mechanisms are needed to optimize screening and risk stratification in this population.

A recent large-scale proteomic study in the general population identified 37 novel heart failure biomarkers that are independent of clinical risk factors, 10 of which are thought to be causal.^21^ Two recent cross-sectional discovery studies of echocardiographic indices identified a signature of subclinical left atrial remodeling that was heightened in adults with HIV compared with persons without HIV, and a 4-protein signature of abnormal myocardial performance index in pediatric PWH.^22,23^ Although protein-based risk prediction may capture HIV disease-specific information beyond traditional and non-traditional risk factors, no studies to date have evaluated proteomic signatures of incident heart failure in a longitudinal cohort of PWH. In the present study, we sought to develop a novel protein-based risk model to predict incident heart failure events among PWH in the Veterans Aging Cohort Study HIV Biomarker Cohort. We hypothesized that protein-based risk models would demonstrate superior performance compared with models based on clinical, inflammatory, or HIV-specific risk factors.

## Methods

### Study population

The Veterans Aging Cohort Study (VACS) HIV is a longitudinal cohort of veterans living with HIV and uninfected veterans matched on age, sex, race-ethnicity, and geographic region that has been previously described.^24^ The VACS Biomarker Cohort (VACS BC) is a subset of VACS HIV, which prospectively enrolled veterans with (n = 1525) and without HIV (n = 843). From 2005 to 2007, VACS BC participants provided informed consent for blood samples. These specimens were collected using a serum separator and EDTA blood collection and shipped to a central repository at the Massachusetts Veterans Epidemiology Research and Information Center (Boston, MA). The date of the blood draw was used as the baseline date for each participant in VACS BC. Participants were followed from their baseline date until death or until censored. The original VACS BC study was approved by the Institutional Review Boards (IRB) at all VACS locations including Yale University (#1506016006) and VA Connecticut Healthcare System (#AJ0013), which included written consent prior to enrollment. As the present study is based on secondary analysis of de-identified data, no additional informed consent or IRB approval was required.

### Data collection and laboratory procedures

Clinical and demographic data closest to the date of blood sample collection were extracted from VACS BC participants’ electronic health records. Variables of interest included age, sex, self-reported race and ethnicity, medical conditions, medication use, smoking status, blood pressure, height and weight, HIV-1 RNA viral load, and CD4^+^ T-cell count, among others. Baseline circulating inflammatory and coagulation markers were previously measured in VACS BC.^25^ Levels of the soluble cluster-of-differentiation 14 (sCD14) protein were measured using ELISA (Quantikine sCD14 immunoassay, R&D Systems, Minneapolis, MN). Interleukin-6 (IL-6) levels were measured using a chemiluminescent immunoassay (QuantiGlo IL-6 immunoassay; R&D Systems, Minneapolis, MN). D-dimer was measured using an immuno-turbidimetric assay (Liatest D-DI; Diagnostica Stago, Parsippany, NJ).

### Proteomic profiling by modified aptamers

We used the SomaScan v4.0 assay (5,284 SOMAmer reagents), a single-stranded DNA aptamer-based platform, to quantify proteins in VACS BC specimens collected at study baseline. Methods for modified aptamer-based protein quantification have been described previously.^26^ The SomaScan platform is an assay based on modified aptamers, which are chemically modified single-strand deoxyribonucleic acid approximately 40 nucleotides in length that act as binding reagents for target proteins. Prior studies assessing fidelity of aptamer-protein binding have demonstrated that >95% of aptamers correctly targeted the intended proteins, provided the proteins are present in sufficient concentration to be detected by mass spectrometry.^26^ Samples are assayed at three different dilutions on the SomaScan platform to ensure each analyte is assayed within its linear range of concentrations. Results are then quantified on a hybridization microarray and reported in relative fluorescence units (RFU). We followed typical procedures for calibration, standardization, and internal controls.^27^ After quality control (QC) filtering performed by SomaLogic (4,268 SOMAmers passing the QC ratio), removal of non-human and control aptamers, and collapsing duplicate reagents, 4,248 protein targets remained for analysis.

### Outcomes

The primary outcome was time to incident heart failure event. As we and others have done previously, heart failure was defined by the presence of at least one inpatient (discharge diagnosis) or at least two outpatient VA, Medicare, or VA fee-for-service International Classification of Diseases (ICD) -9 or ICD-10 codes.^28^ We excluded prevalent cases of heart failure at baseline. We evaluated 5-year heart failure-free survival as a secondary endpoint in our calibration analyses.

### Data processing and statistical analysis

We used descriptive statistics to compare demographic and clinical variables between PLWH who did and did not have an incident heart failure event. We compared continuous variables using two-sample t-tests or Wilcoxon rank-sum tests for mean and median values, respectively. We used Fisher’s exact test to compare binary variables and Chi-squared tests for categorical variables. A p-value of <0.05 was considered statistically significant for our descriptive analyses.

Proteomics data underwent several preprocessing steps, including outlier capping using a 5x median absolute deviation (MAD) threshold (i.e., MAD-based winsorization), followed by log_2_ transformation and median-centered standardization (MAD-scaling). This approach ensured robust normalization and comparability across proteins, reducing the influence of extreme values. The final dataset used for modeling was log2-transformed and MAD-scaled. Clinical heart failure risk factors and HIV-specific variables were incorporated into Cox proportional hazards models for incident heart failure. For risk models involving proteins, we used least absolute shrinkage and selection operator (LASSO) regression with 10-fold cross-validation to identify the optimal set of predictors. The regularization parameter (λ) was selected using the minimum cross-validation partial likelihood deviance. LASSO served as a variable selection tool, shrinking the coefficients of less informative markers to zero while retaining the most predictive markers for inclusion in the final model. For models combining proteins with other clinical or laboratory parameters, we constructed joint predictor pools consisting of the relevant non-protein covariates (e.g., clinical risk factors, CD4^+^ cell count, viral load, inflammatory/coagulation markers) plus the validated proteins, to which we applied LASSO; thus, both protein and non-protein predictors were subjected to penalization in the combined models. Model performance was evaluated using Harrell’s Concordance Index (C-index), which quantifies the discriminative ability of the Cox proportional hazards model. The C-index was computed on the held-out test dataset to assess model fit and predictive accuracy.

For the primary comparisons between the protein-only model and the clinical models, we formally compared C-indices using nonparametric bootstrapping. We generated 1,000 bootstrap resamples of the test dataset, refit both models in each resample, and calculated the distribution of the C-index difference to obtain two-sided p-values and 95% confidence intervals. Other model contrasts were not subjected to formal hypothesis testing. Model calibration was assessed in the test dataset by grouping individuals into quintiles of predicted risk and estimating observed 5-year heart failure-free survival within each quintile using Kaplan-Meier curves. Observed survival estimates (with 95% confidence intervals based on Greenwood’s formula) were plotted against mean predicted survival to generate a quintile-based calibration plot.

### Pathway analysis

We performed exploratory pathway analyses on the subset of proteins that were included as candidate predictors in LASSO-Cox regression models (m = 108). Using their UniProt ID numbers, we used the biomaRt R package to map these to Entrez Gene ID numbers to facilitate the use of pathway enrichment tools.^29–31^ After deduplication, enrichment was performed with clusterProfiler and ReactomePA across Gene Ontology (GO) Biological Process, Kyoto Encyclopedia of Genes and Genomes (KEGG), and Reactome pathways.^32–34^ All analyses used a Benjamini-Hochberg false discovery rate (FDR) correction (q <0.05). Results were exported, sorted by FDR-adjusted p-value, and visualized as dot plots (top 20 categories per database) to illustrate the top pathways and number of key proteins were represented in each.

### Data availability

Due to US Department of Veterans Affairs (VA) regulations and ethics agreements, the analytic data sets used for this study are not permitted to leave the VA firewall without a Data Use Agreement. This limitation is consistent with other studies based on VA data. However, VA data are made freely available to researchers with an approved VA study protocol. For more information, please visit https://www.virec.research.va.gov or contact the VA Information Resource Center at.

### Code availability

Only publicly available statistical packages were used in the analyses for this study as detailed in the Methods section. Statistical code and details of packages employed are available from the corresponding author upon reasonable request.

## Results

### VACS cohort and heart failure outcomes

After excluding baseline prevalent cases of heart failure, a total of 240 incident heart failure events occurred among 1,398 PWH over 13.1 years of median follow-up (2005 to 2019). **Table 1** shows baseline characteristics of the study population, stratified by whether participants did (n = 240) or did not (n = 1158) have an incident heart failure event. The mean age and standard deviation (SD) of PLWH in VACS was 52.1 (SD 8.1) years, and participants with heart failure were older at baseline (51.5 [SD 8.1] vs 54.9 [7.6] years, p <0.001). The baseline prevalence of traditional cardiovascular risk factors was higher among persons with incident heart failure, including hypertension (65% vs 43.4%, p <0.001), diabetes (33.3% vs 14.6%, p <0.001), hyperlipidemia (40.4% vs 30.7%, p = 0.003), and cocaine use (42.9% vs 33.6%, p = 0.006). Circulating levels of soluble CD14 (sCD14) and D-dimer were also higher among those who had an incident heart failure event (p <0.001 and p = 0.006, respectively). Mean systolic blood pressure was also higher among those with an incident heart failure event (130.8 [SD 15.5] vs 127.8 [13.9] mmHg, p = 0.003). There was no significant difference in CD4^+^ T-cell count, HIV viral load, antiretroviral therapy regimen, or cholesterol levels between those who did or did not have an incident heart failure event. There were no significant differences in clinical characteristics between testing and training sets, except for the distribution of female participants (3.6% vs 1.5% in training and testing sets, respectively; p = 0.025; **Supplementary Table S1**).

**Table 1:** Baseline demographic and clinical characteristics among PLWH in the Veterans Aging Cohort Study Biomarker Cohort, stratified by incident heart failure event.

|  | Overall<br>(N=1398) | Incident Heart Failure |  |  |
| --- | --- | --- | --- | --- |
|  |  | No<br>(N=1158) | Yes<br>(N=240) | P-value |
| Mean age, years (SD) | 52.1 (8.1) | 51.5 (8.1) | 54.9 (7.6) | **<0.001** |
| Female, n (%) | 41 (2.9%) | 38 (3.3%) | 3 (1.2%) | 0.090 |
| Race/Ethnicity, n (%) |  |  |  | 0.246 |
| White | 266 (19.0%) | 216 (18.7%) | 50 (20.8%) |  |
| Black | 960 (68.7%) | 805 (69.5%) | 155 (64.6%) |  |
| Hispanic | 118 (8.4%) | 97 (8.4%) | 21 (8.8%) |  |
| Other | 54 (3.9%) | 40 (3.5%) | 14 (5.8%) |  |
| Medical history, n (%) |  |  |  |  |
| Hypertension | 659 (47.1%) | 503 (43.4%) | 156 (65.0%) | **<0.001** |
| On antihypertensive treatment | 952 (68.1%) | 759 (65.5%) | 193 (80.4%) | <0.001 |
| Diabetes | 249 (17.8%) | 169 (14.6%) | 80 (33.3%) | **<0.001** |
| Hyperlipidemia | 452 (32.3%) | 355 (30.7%) | 97 (40.4%) | **0.003** |
| On lipid-lowering therapy | 402 (28.8%) | 317 (27.4%) | 85 (35.4%) | 0.012 |
| Overweight/obese (body mass index >25 kg/m<sup>2</sup>) | 736 (52.8%) | 604 (52.3%) | 132 (55.0%) | 0.453 |
| Current smoker | 689 (49.3%) | 557 (48.1%) | 132 (55.0%) | 0.052 |
| Cocaine or crack use | 294 (22.1%) | 233 (21.1%) | 61 (27.2%) | 0.120 |
| Methamphetamine and other stimulant use | 59 (4.5%) | 50 (4.5%) | 9 (4.1%) | 0.820 |
| Prior ASCVD event | 69 (4.9%) | 45 (3.9%) | 24 (10.0%) | **<0.001** |
| Mean (SD) systolic blood pressure, mmHg | 128.3 (14.2) | 127.8 (13.9) | 130.8 (15.5) | **0.003** |
| Mean (SD) body mass index, kg/m<sup>2</sup> | 25.9 (4.8) | 25.8 (4.7) | 26.2 (5.5) | 0.261 |
| On antiretroviral regimen at baseline, n (%) | 1144 (81.8%) | 943 (81.4%) | 201 (83.8%) | 0.397 |
| NRTI | 1156 (82.7%) | 950 (82.0%) | 206 (85.8%) | 0.157 |
| NNRTI | 499 (35.7%) | 418 (36.1%) | 81 (33.8%) | 0.490 |
| Mean (SD) laboratory values |  |  |  |  |
| Total Cholesterol, mg/dL | 176.6 (43.1) | 177.2 (44.2) | 173.7 (37.3) | 0.259 |
| LDL Cholesterol, mg/dL | 99.9 (34.6) | 100.4 (34.8) | 97.4 (34.0) | 0.244 |
| HDL Cholesterol, mg/dL | 44.5 (16.8) | 44.6 (16.6) | 43.9 (17.3) | 0.548 |
| Estimated glomerular filtration rate, mL/min/1.73 m<sup>2</sup> | 99.0 (30.4) | 99.7 (28.9) | 95.6 (36.8) | 0.054 |
| CD4<sup>+</sup> count, cells/mm<sup>3</sup> | 446 (286) | 451 (289) | 417 (270) | 0.090 |
| Nadir CD4<sup>+</sup> count, cells/mm<sup>3</sup> | 229 (178) | 232 (179) | 218 (175) | 0.269 |
| HIV viral load, copies/mL | 20623 (75828) | 20441 (77896) | 21499 (65085) | 0.844 |
| Soluble CD14 (sCD14), ng/mL | 1810 (543) | 1777 (529) | 1971 (579) | **<0.001** |
| D-dimer, ng/mL | 0.5 (1.0) | 0.4 (0.7) | 0.6 (1.8) | **0.006** |
| Interleukin-6 (IL-6), pg/mL | 3.2 (7.5) | 3.2 (7.7) | 3.6 (6.4) | 0.425 |

### Development and performance of clinical, inflammatory, and protein risk models

The sample was divided randomly 2:1 into training (n = 932) and test (n = 466) sets, each of which included 164 and 76 participants with incident heart failure events, respectively. We developed clinical, inflammatory marker, and protein-based risk models in the training set and evaluated model fit in the test set (**Table 2**). Among the clinical models, the ASCVD 10-year risk score by the Pooled Cohort Equations (Model 1) demonstrated a test C-index of 0.607 (95% confidence interval [CI] 0.544 – 0.670), while individual clinical heart failure risk factors (Model 2; age, sex, race, systolic blood pressure, antihypertensive use, diabetes, smoking status, body mass index, total cholesterol, high-density lipoprotein cholesterol) yielded a C-index of 0.639 (0.574 – 0.704). Among the clinical risk factor models, individual traditional risk factors in combination with HIV disease markers (CD4^+^ T-cell count, viral load) exhibited the best predictive performance (Model 5b; C-index 0.656 [95% CI 0.591 – 0.720]). Addition of circulating inflammatory and coagulation markers (soluble CD-14 [sCD14], interleukin-6 [IL-6], D-dimer) did not substantially improve performance (Model 8; C-index 0.657 [0.594 – 0.720]).

**Table 2:** Predictive performance of multivariable models for incident heart failure among PLWH.

| Model | Covariates included in model | Proteins selected in model | Event-free survival C-index<br>(95% confidence interval) |  |
| --- | --- | --- | --- | --- |
|  |  |  | Training set<br>(n = 932) | Test set<br>(n = 466) |
| CLINICAL RISK MODELS |  |  |  |  |
| 1 | ASCVD 10-year risk score<sup>1</sup> | -- | 0.681 (0.640 - 0.722) | 0.607 (0.544 - 0.670) |
| 2 | Clinical heart failure risk factors<sup>2</sup> | -- | 0.724 (0.687 - 0.761) | 0.639 (0.574 - 0.704) |
| 3a | Model 1 + eGFR<sup>3</sup> | -- | 0.661 (0.616 - 0.706) | 0.607 (0.544 - 0.670) |
| 3b | Model 2 + eGFR | -- | 0.724 (0.685 - 0.763) | 0.640 (0.575 - 0.705) |
| 4a | Model 1 + eGFR + stimulant use | -- | 0.685 (0.640 - 0.730) | 0.630 (0.567 - 0.693) |
| 4b | Model 2 + eGFR + stimulant use | -- | 0.738 (0.699 - 0.777) | 0.647 (0.580 - 0.714) |
| 5a | Model 3a + CD4<sup>+</sup> count + viral load | -- | 0.707 (0.666 - 0.748) | 0.637 (0.576 - 0.698) |
| 5b | Model 3b + CD4<sup>+</sup> count + viral load | -- | 0.754 (0.717 - 0.791) | 0.656 (0.591 - 0.721) |
| INFLAMMATORY MARKER RISK MODELS |  |  |  |  |
| 6 | sCD14 + IL-6 + D-dimer | -- | 0.638 (0.593 - 0.683) | 0.611 (0.542 - 0.680) |
| 7 | ASCVD score + sCD14 + IL-6 + D-dimer | -- | 0.711 (0.672 - 0.750) | 0.632 (0.567 - 0.697) |
| 8 | Clinical heart failure risk factors<sup>2</sup> + sCD14 + IL-6 + D-dimer | -- | 0.749 (0.712 - 0.786) | 0.645 (0.580 - 0.710) |
| 9 | Model 6 + CD4<sup>+</sup> count + viral load | -- | 0.641 (0.598 - 0.684) | 0.600 (0.529 - 0.671) |
| 10 | Model 7 + CD4<sup>+</sup> count + viral load |  | 0.720 (0.683 - 0.757) | 0.632 (0.567 - 0.697) |
| 11 | Model 8 + CD4<sup>+</sup> count + viral load |  | 0.757 (0.722 - 0.792) | 0.657 (0.594 - 0.720) |
| PROTEIN RISK MODELS |  |  |  |  |
| 12 | ASCVD score + **proteins** | RNASE1, NBL1, HE4, GAS1, FBLN3, TAGLN, FBLN5, TIM-1 | 0.740 (0.651 - 0.812) | 0.713 (0.582 - 0.815) |
| 13 | Clinical heart failure risk factors<sup>2</sup> + **proteins** | RNASE1, HE4, GAS1, TAGLN, FBLN5, TIM-1 | 0.753 (0.666 - 0.824) | 0.717 (0.587 - 0.819) |
| 14 | ASCVD score + sCD14 + D-dimer + IL-6 + **proteins** | RNASE1, NBL1, HE4, GAS1, TAGLN, FBLN5, TIM-1 | 0.740 (0.651 - 0.812) | 0.710 (0.579 - 0.814) |
| 15 | **Proteins only** | RNASE1, HE4, TAGLN, FBLN5, TIM-1, FABPA, MMP-12, GPNMB | 0.719 (0.632 - 0.792) | 0.725 (0.597 - 0.825) |
| 16 | **Proteins** + CD4<sup>+</sup> count + viral load | RNASE1, HE4, PXDN, TAGLN, FBLN5, TIM-1, GPNMB | 0.716 (0.629 - 0.789) | 0.715 (0.587 - 0.816) |
| 17 | **Proteins** + CD4<sup>+</sup> count + viral load + sCD14 + IL-6 + D-dimer | RNASE1, HE4, PXDN, TAGLN, FBLN5, TIM-1, GPNMB | 0.714 (0.627 - 0.789) | 0.720 (0.591 - 0.820) |

<sup>1</sup> ACC/AHA Pooled Cohort Equations
<sup>2</sup> Age; sex; race; systolic blood pressure; antihypertensive medication use; diabetes; current smoker; body mass index; total cholesterol; high-density lipoprotein cholesterol
<sup>3</sup> eGFR, estimated glomerular filtration rate
<sup>4</sup> sCD14, soluble CD14; IL-6, interleukin-6
C-index

**Table 3:**
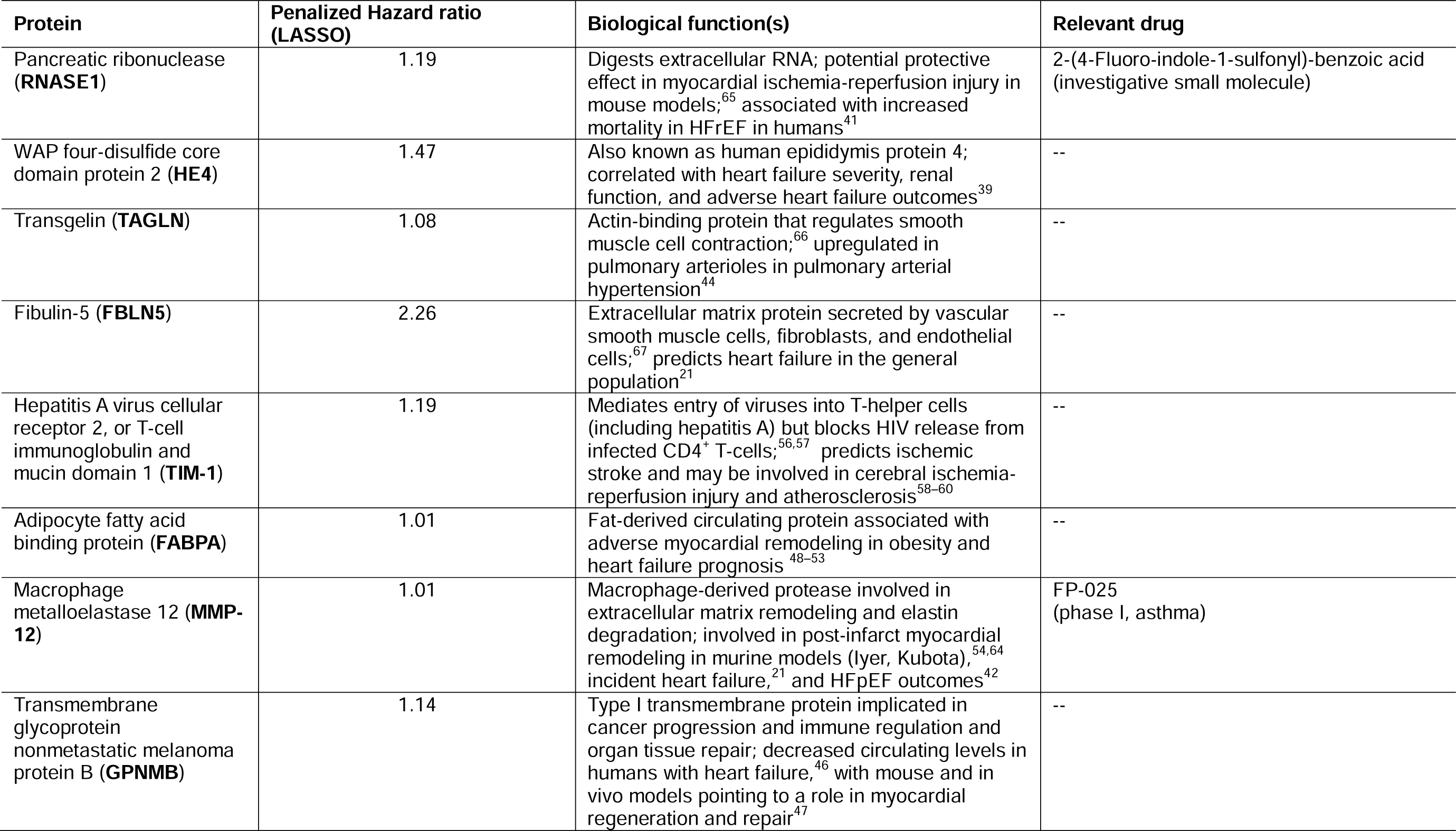
Major functions of individual proteins in the final 8-protein incident heart failure model.

To develop protein risk models, we initially screened all 4,248 proteins in the training set in univariable single-protein Cox proportional hazards models of incident heart failure. After stringent Bonferroni correction (p <1E-5), we identified 227 of 4,248 proteins that predicted incident heart failure; these 227 were then validated in the test set in univariable single-protein Cox models. This two-step selection process yielded 108 of 227 candidate proteins that passed the second Bonferroni threshold (p <2E-4). These 108 proteins were included as candidate predictors for downstream multivariable LASSO-Cox regression models (**Figure 1**).

**Figure 1:**
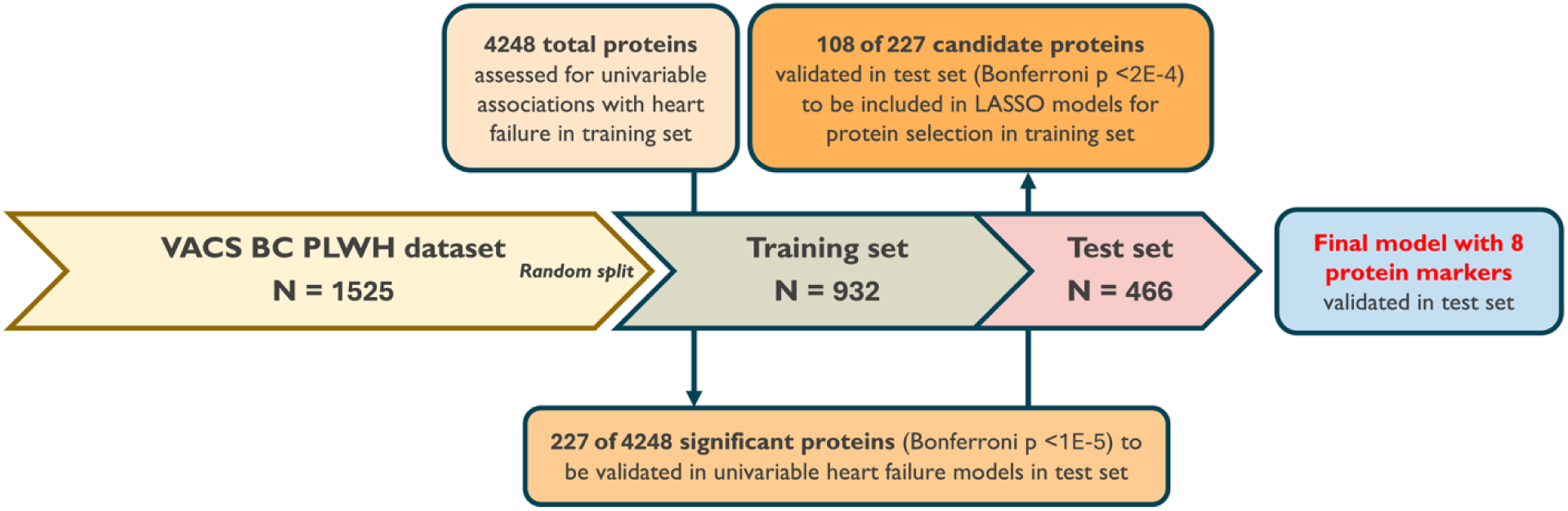
Candidate protein selection process for inclusion in incident heart failure models.

LASSO-Cox regression identified a subset of 8 protein markers in a protein-only risk model of incident heart failure (Model 15): pancreatic ribonuclease (RNASE1), WAP four-disulfide core domain protein 2 (HE4), transgelin (TAGLN), fibulin 5 (FBLN5), hepatitis A viral cellular receptor 1, also known as T-cell immunoglobulin and mucin domain 1 (TIM-1), adipocyte fatty acid-binding protein (FABPA), macrophage metalloelastase (MMP-12), and transmembrane glycoprotein nonmetastatic melanoma protein B (GPNMB; **Tables 2 & 3, Figure 2**). Plasma concentrations of all proteins differed between PWH who did and did not have an incident heart failure event (p <0.001 for all; **Supplementary Table S2**). The protein-only model (Model 15) outperformed the clinical risk model (Model 2) in prediction of incident heart failure events in the test set (C-indices 0.725 [95% CI 0.597 – 0.825] vs 0.639 [0.574 – 0.704]; p = 0.033). In combined models, LASSO-Cox regression selected between 6 and 8 proteins in addition to clinical and/or inflammatory markers, all of which included RNASE1, TAGLN, and FBLN5. C-indices for all models incorporating protein markers were ≥0.710 (**Table 2**); the protein-only model (Model 15) exhibited the highest C-index among all the combined protein models. There was no meaningful improvement in model performance when inflammatory markers and HIV-specific variables (sCD14, IL-6, D-dimer, CD4+ count, viral load) were added to the protein model (Model 17; C-index 0.720 [0.591 – 0.820]; p = 0.0592 vs Model 15). In the LASSO selection process, none of these variables were retained in the final penalized model, indicating limited incremental predictive value beyond the protein signature.

**Figure 2:**
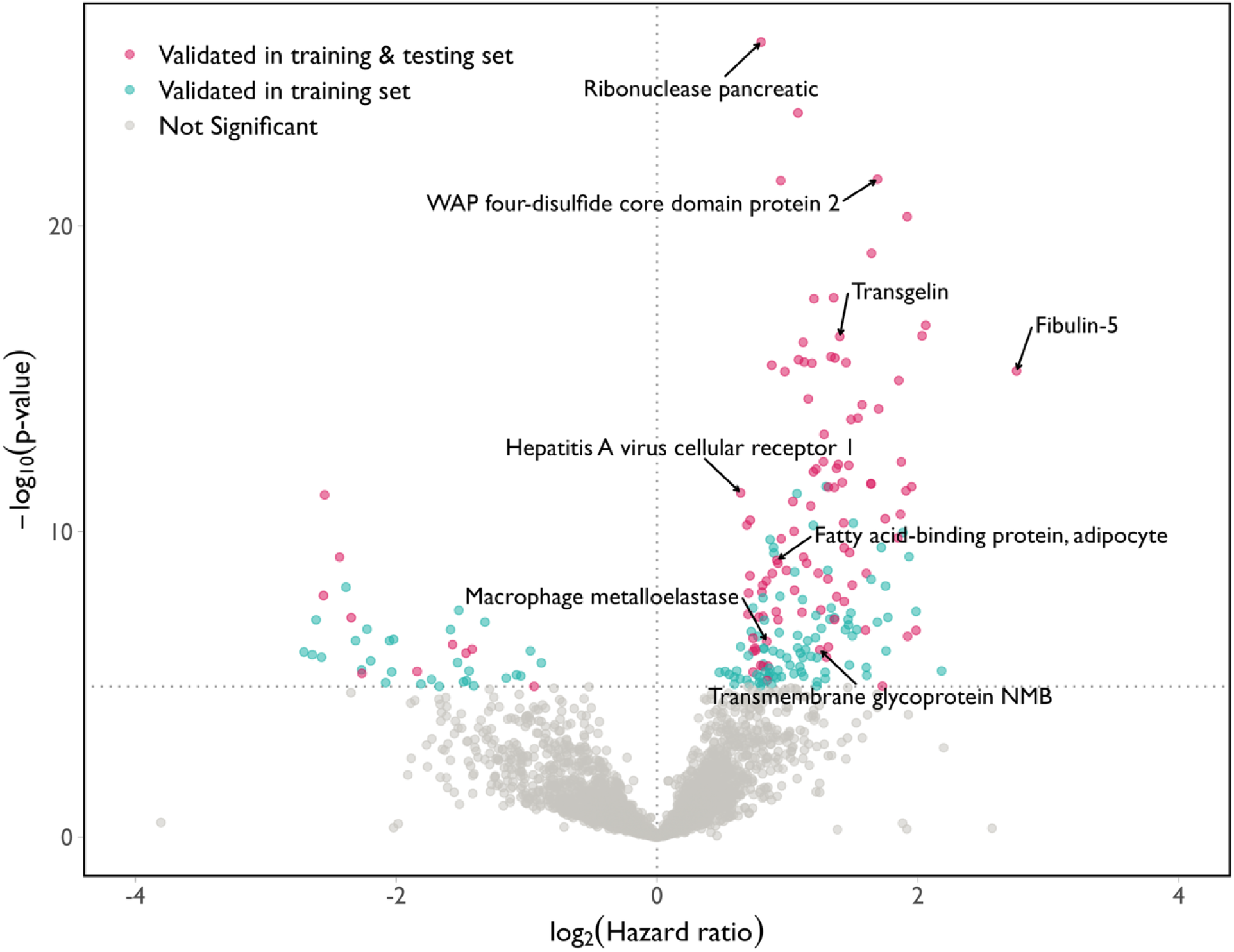
Proteins in sequential phases of selection and model development. Horizontal and vertical dotted lines represent first-round Bonferroni-corrected significance threshold (p <1E-5) and hazard ratio of 1.0, respectively. Markers selected in final protein-only model of incident heart failure are labeled.

We compared heart failure event-free survival in the test set (n = 466) stratified into risk quartiles using Model 2 (clinical risk factors) and Model 15 (proteins only; **Figure 3**). The clinical risk factor model did not discriminate between risk quartiles in terms of incident heart failure risk (p = 0.97), while the protein-only model demonstrated divergence between risk quartiles within the first 5 years of follow-up (p <0.0001). Participants in the highest-risk quartile based on the protein model had a Kaplan-Meier-estimated 10-year cumulative incidence of heart failure of approximately 33%, whereas the incidence in the lowest-risk quartile was approximately 2%, demonstrating the broad dynamic range of the protein model.

**Figure 3:**
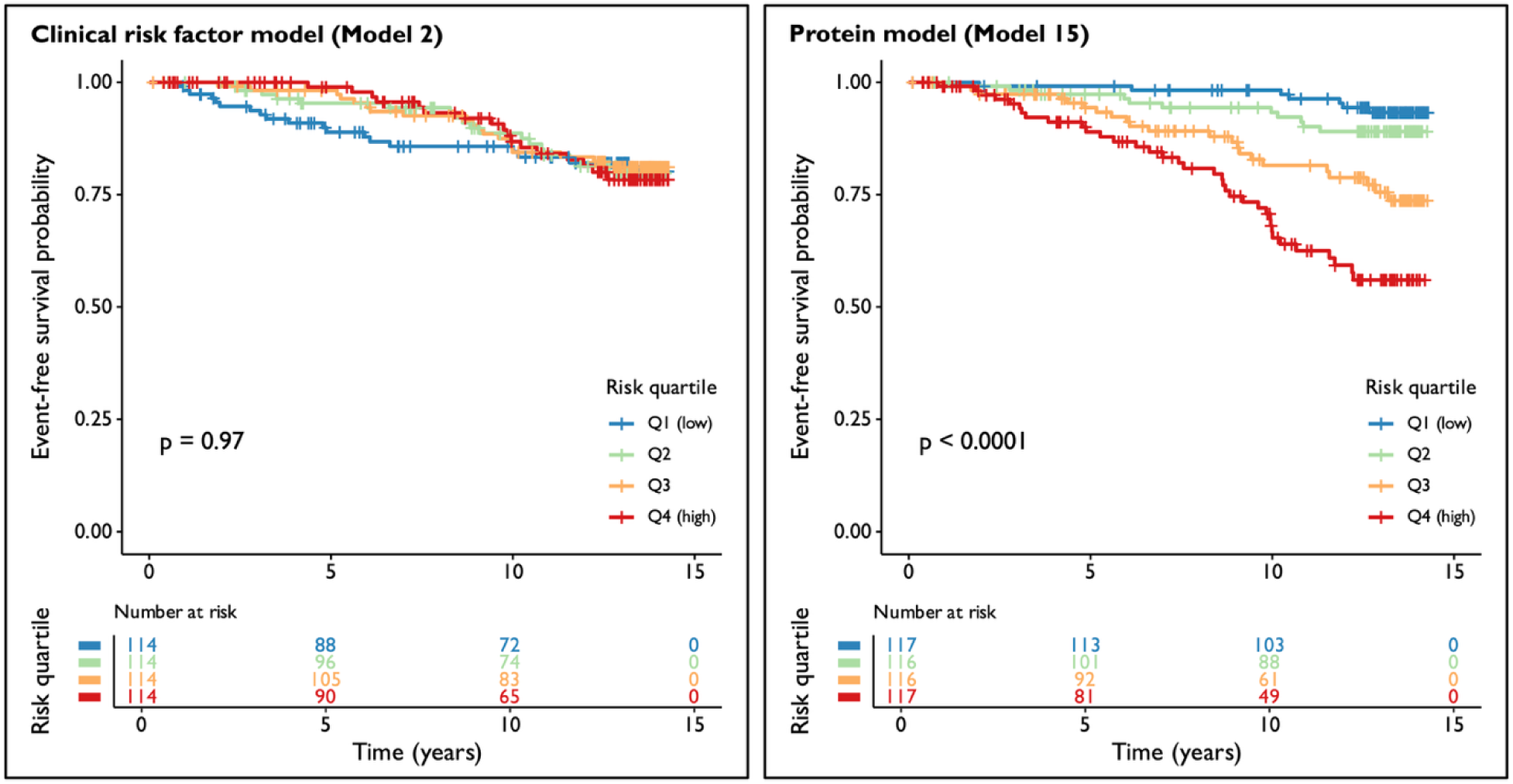
Quartile-based Kaplan-Meier curves for incident heart failure-free survival in test set of PWH (n = 466). Individuals were stratified into predicted risk quartiles based on models incorporating clinical risk factors (Model 2) and proteins only (Model 15). The clinical model included 455 total participants, as 10 participants were missing one or more clinical risk factor parameters, and thus could not be risk-stratified. By contrast, protein data was available for all 466 participants. The protein-only model demonstrated improved risk discrimination compared with the clinical model, particularly in the highest-risk quintile.

As a validation exercise, we fit models derived in the PWH training set to VACS participants without HIV (PWoH; n = 715). Their baseline clinical and demographic characteristics are provided in **Supplementary Table S3**. Over 12.9 years of median follow-up, there were 143 incident heart failure events among 715 PWoH in VACS. Model performance in PWoH is summarized in **Supplementary Table S4**. PWH-derived models that included clinical risk scores or individual clinical risk factors performed better in PWoH. The PWH-derived inflammatory marker model (Model 6) performed similarly in PWoH as in PWH (C-indices: 0.589 [95% CI 0.542 – 0.636] vs 0.611 [0.542 – 0.680], respectively). Among the PWH-derived protein-based models, the combined clinical risk factor and protein model (Model 13) had the highest C-index (0.730 [0.639 – 0.806]). This combined PWH-derived model selected 6 proteins, of which 5 were shared with the PWH-derived protein-only model (RNASE1, HE4, TAGLN, FBLN5, TIM-1), with the addition of growth arrest-specific protein 1 (GAS1). The PWH-derived protein-only model (Model 15) had the lowest C-index in PWoH (0.699 [0.608 – 0.777]) among the protein models.

### Model calibration

Figure 4 shows a quintile-based calibration curve for the 8-protein incident heart failure risk model in the test set (n = 466). A total of 76 incident heart failure events occurred in the test cohort over the full follow-up period; calibration was evaluated specifically at the 5-year time horizon using Kaplan-Meier estimates. Predicted and observed 5-year survival were closely aligned across quintiles, with overlapping 95% confidence intervals and no evidence of systematic over- or under-prediction, indicating reliable model calibration.

**Figure 4:**
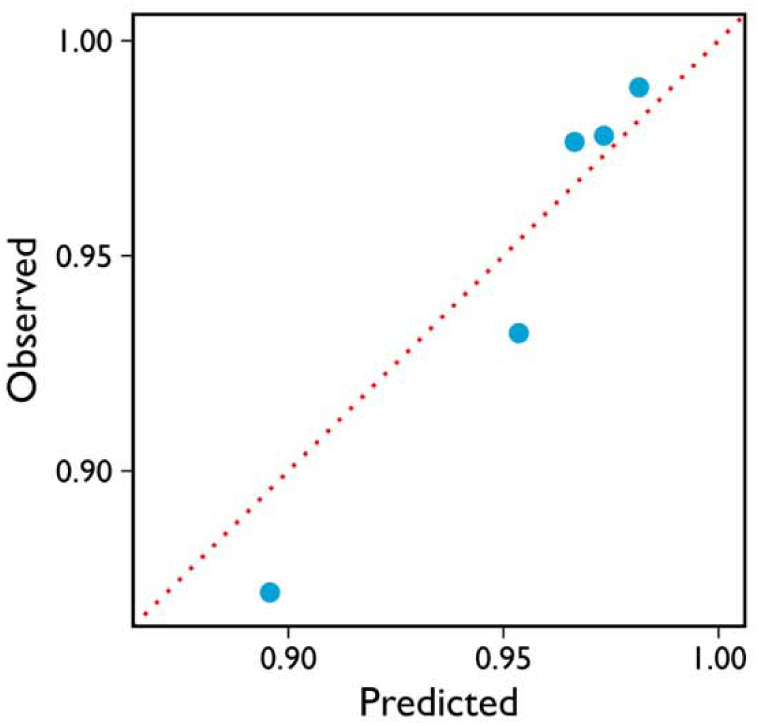
Quintile-based calibration curve in test set (n = 466) for observed vs predicted 5-year heart failure-free survival based on protein-only model. All 95% confidence intervals for each point (not shown) overlap with the dashed red line, which represents perfect calibration between observed and predicted events.

### Protein correlations with clinical risk factors and exploratory pathway analysis

Figure 5 shows the correlation matrix between the 8 proteins selected in the final model and established clinical heart failure risk factors, with the corresponding hazard ratio (HR) for each protein shown at the bottom. Positive correlations (purple) indicate that higher protein levels were associated with higher values of the corresponding clinical variable, whereas negative correlations (green) indicate inverse associations. The HR color scale illustrates each protein’s effect size on incident heart failure in the test set. Correlations between protein levels and clinical risk factors were generally modest (| r | <0.3), suggesting that these biomarkers capture complementary information beyond that afforded solely by clinical risk factors.

**Figure 5:**
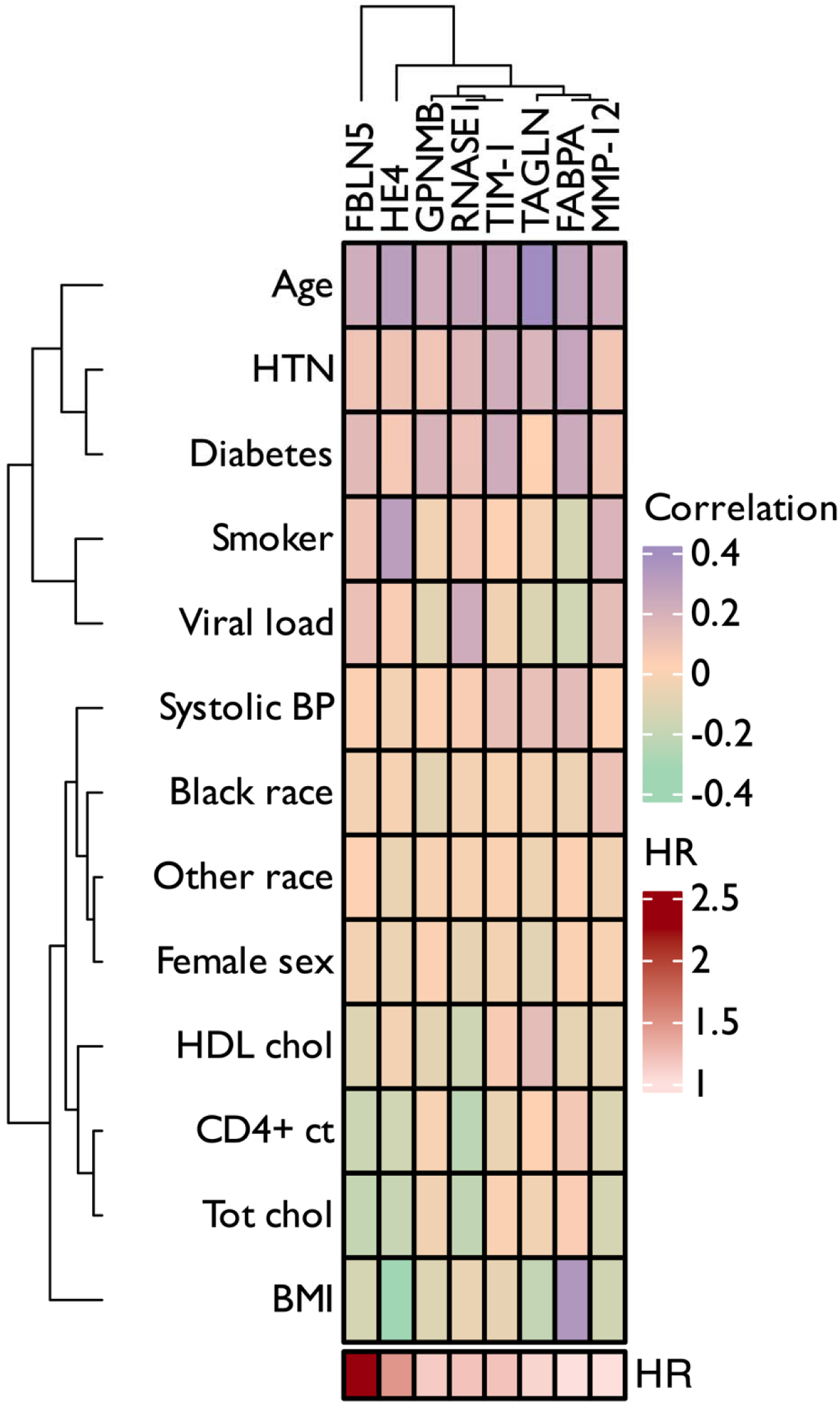
Correlation matrix between clinical risk factors and final 8 selected protein markers in incident heart failure models. Proteins are ordered by hierarchical clustering of their penalized hazard ratios, and clinical variables are hierarchically clustered based on similarity of their correlation profiles across the 8 proteins, thereby grouping variables with similar correlation patterns.

To gain potential mechanistic insights into protein networks involved in incident heart failure, we performed an exploratory pathway enrichment analysis. We based this analysis on the 108 candidate markers for LASSO-Cox regression models from the 4,248 background proteins. This analysis demonstrated significant enrichment for proteins involved in a variety of pathways, including cytokine/immunoregulatory signaling and cellular response to viruses; cellular programming and intracellular signaling; chemotaxis; and extracellular matrix organization (**Supplementary Figures S1-S3**).

The full list of proteins significantly associated with incident heart failure validated in the test set (m = 108) and included as candidates in the predictive protein models is presented in **Supplementary Table S5**. Among these, 44 were druggable targets whose protein structures have been delineated and published based on the Therapeutic Target Database.^35^

## Discussion

In this study of proteomic signatures of heart failure in PWH, we quantified 4,248 plasma proteins in a prospective longitudinal cohort of United States Veterans with and without HIV. We identified an 8-protein plasma signature at baseline that predicted incident heart failure in PWH and outperformed traditional, inflammatory, and HIV-specific clinical risk models. Our protein model was well-calibrated and correlations between individual proteins and risk factors were modest, suggesting that the selected protein biomarkers capture biology that is largely independent of established clinical heart failure risk factors and HIV-specific parameters. Among PWH with heart failure, we noted upregulated pathways involved in immune and intracellular signaling, cellular programming, chemotaxis, and extracellular matrix organization, among others. Our findings point to potential biological pathways of heart failure in HIV that can be leveraged for risk stratification and as potential treatment targets in this population.

### Protein markers predict clinical heart failure in the general population

To our knowledge, this is the first large-scale proteomic profiling study of incident heart failure in a longitudinal cohort of persons living with HIV. Notably, despite deriving models in PWH, 6 of the 8 proteins in our final model have been described in association with clinical heart failure in the general population without HIV. These prior studies evaluated development of incident heart failure and prognosis in patients with existing heart failure. Three markers—FBLN5, MMP-12, and TAGLN—predicted incident heart failure in a recent proteomic profiling analysis in the general population.^21^ Similarly, in patients with chronic kidney disease, another large-scale analysis externally validated three of our markers (FBLN5, TAGLN, HE4) in incident heart failure; in a separate targeted study, HE4 predicted death and readmission in patients with chronic kidney disease hospitalized with acute heart failure.^36,37^ Five of the markers we identified have been associated with increased mortality in heart failure with reduced ejection fraction (FABPA, HE4, RNASE1, TAGLN) and heart failure with preserved ejection fraction (FABPA, HE4, MMP-12) in the general population.^38–42^ While we derived our final protein model exclusively in PWH, taken together, our results are reinforced by other studies reporting several of the same proteomic markers of clinical heart failure in the general population. Further, the consistency of the results despite the various proteomic platforms used throughout the literature points toward a true association of these proteins with clinical heart failure.

### Protein markers of subclinical myocardial remodeling in HIV

None of our final markers of incident heart failure in PWH overlapped with two recent discovery investigations of echocardiographic myocardial remodeling in adults and children with HIV, though proteins identified by both studies have been linked with cardiovascular disease in persons without HIV.^22,23^ HIV is an independent risk factor for myocardial disease; while unique HIV-specific mechanisms are thought to mediate this risk, the overlap in our PWH-derived heart failure markers with those published in the general population suggests shared biology of heart failure between PWH and the general population.^2^ The consistency of predictive markers across various studies, proteomic platforms, and subpopulations indicates that clinical heart failure pathology favors a final common pathway—regardless of etiology, HIV, or systemic disease substrate—in which these proteins play a biological role and/or reflect heart failure physiology. Indeed, HIV may accelerate or amplify these core heart failure pathways, or act further upstream via HIV-specific mechanisms; this is further supported by the lack of strong correlation between our final proteins and clinical risk factors, including HIV-specific parameters. Similarly, our PWH-derived protein model did not perform as well when fit to VACS BC participants without HIV. While the selected protein markers may be common to both persons with and without HIV, their weighting coefficients likely differ, thus driving the relatively poorer fit in those without HIV. This highlights a limitation in our sample size, as the number of VACS BC participants without HIV was insufficient to support independent model derivation and validation. Further, we used a stringent two-step Bonferroni correction to identify the candidate pool of protein markers for our final models, which may have screened out important markers through an overly conservative selection process.

### Potential biological functions of identified proteins and implications in HIV-specific pathways

While our exploratory analyses identified several important pathways upregulated in heart failure, the exact biological role of our final 8 predictors in PWH remains unclear. In addition to clinical prognostication, several of these markers have been associated with structural and functional myocardial abnormalities, including FBLN5 with right ventricular dysfunction, TAGLN with pulmonary arteriolar smooth muscle cell proliferation in congenital heart disease-related pulmonary hypertension, MMP-12 and GPNMB with post-infarct myocardial remodeling, and FABPA with subclinical myocardial abnormalities in obesity and its associated metabolic complications.^39,43–54^

Whether these markers have additional specificity or a role in HIV-associated heart failure pathogenesis remains to be determined. Circulating FABPA, for instance, did not differ by HIV status in one study, but was associated with age, fat composition, BMI, lipids, and cardiovascular risk.^55^ Similarly, hepatitis A viral cellular receptor 1, also known as T-cell immunoglobulin and mucin domain 1 (TIM-1), belongs to the family of cell surface glycoproteins that mediate entry of various viruses into T-helper cells, including hepatitis A. Notably, TIM-1 has been shown to block HIV release from infected CD4^+^ T-cells.^56,57^ TIM-1 has been implicated in incident ischemic stroke in a Swedish cohort without HIV, with genetic analyses pointing to a potential causal role.^58^ Murine studies suggest TIM-1 has a role in cerebral ischemia-reperfusion injury and atherosclerosis, potentially driven by an imbalance in the Th1/Th2 cell ratio.^59,60^ Prospective studies in PWH also signal that higher CD4^+^ Th1 proportion may be associated with incident heart failure.^61^ Whether TIM-1’s potential role in heart failure is mediated via inflammation, and whether HIV-specific mechanisms interact with this pathway, remain to be characterized.

Further mechanistic studies on the heart failure markers we identified can also clarify the directionality of risk. Of our 8 final markers, GPNMB was the only one that has not been validated in proteome-wide outcomes studies of clinical heart failure. However, our finding that GPNMB confers increased risk conflicts with clinical studies demonstrating lower circulating levels in patients with heart failure compared to heart failure-free controls, and with other translational studies pointing to its possible role in myocardial regeneration and repair post-infarction.^46^ A recent comprehensive translational study found that GPNMB expression is increased in failing human hearts and post-infarct murine hearts; its deficiency led to rapid post-infarct left ventricular dysfunction, cardiac rupture, and increased mortality. Conversely, the same study noted that by increasing circulating GPNMB levels, there was enhanced myocyte contraction, reduced fibroblast activation, and improved post-infarct myocardial function.^47^ We also noted an increased risk of heart failure associated with MMP-12, consistent with another large-scale proteomics analysis in the general population and in line with other clinical and translational studies demonstrating association with atherosclerosis development and incident coronary events.^21,62,63^ This is in contrast with murine models showing that MMP-12 may have a role in post-infarct myocardial recovery by inhibiting neutrophil infiltration.^54,64^ In aggregate, these observations suggest that rather than having causal or harmful functions predisposing to heart failure, GPNMB and MMP-12 are upregulated due to their potential protective role in myocardial tissue repair. Further mechanistic and translational studies are needed to clarify these pathways, particularly if their pharmacologic amplification could improve heart failure outcomes in PWH.

### Risk prediction and implications for PWH

While the exact biological roles of our final proteins are not yet defined, our protein model outperformed clinical models in heart failure risk prediction. The broader implications for PWH are significant, especially as atherosclerotic cardiovascular risk prediction algorithms developed in the general population translate poorly in PWH.^18–20^ Further, all-cause mortality in heart failure is higher among PWH than their peers without HIV, highlighting the need for tailored risk stratification tools in this population.^15^ In our study, a proteomic signature measured at a single baseline time point effectively predicted incident heart failure events over long-term follow-up, outperforming clinical and HIV-specific risk models. This is in line with other proteomic profiling studies of heart failure in special populations, including a recent study in patients with chronic kidney disease, which showed that protein risk prediction models consistently outperformed the 2019 Pooled Cohort Equations to Prevent Heart Failure (PCP-HF).^16,36^ Importantly, the input range of clinical parameters for PCP-HF and the 2024 American Heart Association Predicting Risk of CVD EVENTs (PREVENT) Heart Failure equations precluded estimation of risk for up to a quarter of our VACS BC participants; the most commonly out-of-range input variables were body mass index and total cholesterol. While this highlights a limitation of our approach—inability to evaluate performance of contemporary heart failure risk prediction algorithms in comparison to our protein models—the narrow range of inputs likely limits the applicability of these tools to many populations even beyond PWH and lends further support for a protein-based approach.

## Limitations

Despite the strength of our sample and the rigorous study design, our findings must be interpreted in the context of several limitations. Firstly, this was a cohort composed of US Veterans, and consequently primarily male. This raises a second limitation—the lack of an external validation cohort—to assess the generalizability of our results to broader populations of PWH, including females. We also lacked statistical power to examine subtypes of heart failure, such as heart failure with reduced and preserved ejection fraction, individually.

## Conclusions

In summary, we derived a baseline 8-protein plasma signature in PWH that predicted incident heart failure events over 13 years of median follow-up, outperforming clinical, HIV-specific, and inflammatory/coagulation markers. These proteins have been described in relation to heart failure or myocardial remodeling in persons without HIV, suggesting that their biological role may be core to heart failure physiology, regardless of HIV status. Pathways of extracellular matrix organization and cellular and immune signaling were upregulated in our exploratory analysis of 108 candidate markers, but the exact biology of the final 8 protein markers in our predictive model and their mechanistic role in heart failure remain to be determined. To our knowledge, this is the first study to report proteomic markers of incident heart failure in a longitudinal cohort of PWH. While further validation in external cohorts is needed, protein prediction can be used to identify PWH at highest risk for heart failure and druggable targets can be leveraged to develop novel therapeutic targets with the ultimate goal of preventing clinical disease. Future translational research should aim to bridge clinical risk markers with mechanistic pathways underlying heart failure in this population.

## Data Availability

Data used in the present study can be requested from the Veterans Aging Cohort Study (VACS) Consortium.

## Acknowledgments

The authors acknowledge the veterans who participated in the Veterans Aging Cohort Study and the study coordinators and staff at each respective enrollment and coordination sites, without whom this research would not be possible.

## Author contributions

SSS, PYH, MF, and PG designed the study. PG supervised the laboratory work and quality control of the proteomics assays. MF and KS provided access to the clinical data. TG performed the data analysis. SSS drafted the manuscript. All authors reviewed and critically revised the final manuscript and are accountable for all aspects of the work presented.

## Disclaimer

The views and opinions expressed in this manuscript are those of the authors and do not necessarily represent those of the Department of Veterans Affairs nor the United States government.

