## Supplement for "A novel plasma proteomic signature predicts long-term incident heart failure risk among persons living with HIV"

**Supplementary Table S1: Baseline demographic and clinical characteristics among PLWH in the Veterans Aging Cohort Study Biomarker Cohort, stratified by training vs testing set.**

| | | Split | | |
|---|---|---|---|---|
| | **Overall** | **Training set** | **Test set** | **P-value** |
| | **(N=1398)** | **(N=932)** | **(N=466)** | |
| **Mean age, years (SD)** | 52.1 (8.1) | 52.1 (8.1) | 52.1 (8.1) | 0.893 |
| **Female, n (%)** | 41 (2.9%) | 34 (3.6%) | 7 (1.5%) | **0.025** |
| **Race/Ethnicity, n (%)** | | | | 0.598 |
| **White** | 266 (19.0%) | 173 (18.6%) | 93 (20.0%) | |
| **Black** | 960 (68.7%) | 638 (68.5%) | 322 (69.1%) | |
| **Hispanic** | 118 (8.4%) | 81 (8.7%) | 37 (7.9%) | |
| **Other** | 54 (3.9%) | 40 (4.3%) | 14 (3.0%) | |
| **Medical history, n (%)** | | | | |
| **Hypertension** | 659 (47.1%) | 444 (47.6%) | 215 (46.1%) | 0.596 |
| **On antihypertensive treatment** | 952 (68.1%) | 633 (67.9%) | 319 (68.5%) | 0.856 |
| **Diabetes** | 249 (17.8%) | 157 (16.8%) | 92 (19.7%) | 0.182 |
| **Hyperlipidemia** | 452 (32.3%) | 303 (32.5%) | 149 (32.0%) | 0.840 |
| **On lipid-lowering therapy** | 402 (28.8%) | 272 (29.2%) | 130 (27.9%) | 0.659 |
| **Overweight/obese (BMI >25 kg/m$^2$)** | 736 (52.8%) | 489 (52.6%) | 247 (53.2%) | 0.818 |
| **Current Smoker** | 689 (49.3%) | 455 (48.8%) | 234 (50.2%) | 0.623 |
| **Cocaine use** | 294 (22.1%) | 204 (23.0%) | 90 (20.3%) | 0.310 |
| **Methamphetamine and other stimulant use** | 59 (4.5%) | 38 (4.3%) | 21 (4.7%) | 0.810 |
| **Prior ASCVD event** | 69 (4.9%) | 41 (4.4%) | 28 (6.0%) | 0.193 |
| **Mean (SD) systolic blood pressure, mmHg** | 128.3 (14.2) | 127.9 (14.6) | 128.9 (13.4) | 0.225 |
| **Mean (SD) body mass index, kg/m$^2$** | 25.9 (4.8) | 25.8 (4.5) | 26.1 (5.3) | 0.302 |
| **On antiretroviral regimen at baseline, n (%)** | 1144 (81.8%) | 762 (81.8%) | 382 (82.0%) | 0.922 |
| **NRTI** | 1156 (82.7%) | 766 (82.2%) | 390 (83.7%) | 0.484 |
| **NNRTI** | 499 (35.7%) | 334 (35.8%) | 165 (35.4%) | 0.875 |
| **Mean (SD) laboratory values)** | | | | |
| **Total Cholesterol, mg/dL** | 176.6 (43.1) | 177.4 (44.3) | 175.0 (40.6) | 0.335 |
| **LDL Cholesterol, mg/dL** | 99.9 (34.6) | 99.5 (34.6) | 100.6 (34.7) | 0.593 |
| **HDL Cholesterol, mg/dL** | 44.5 (16.8) | 44.2 (16.6) | 45.1 (17.1) | 0.360 |
| **Estimated glomerular filtration rate, mL/min/1.73 m$^2$** | 99.0 (30.4) | 98.1 (29.1) | 100.7 (32.8) | 0.128 |
| **CD4$^+$ count, cells/mm$^3$** | 445.5 (286.2) | 449.6 (289.3) | 437.4 (280.0) | 0.452 |
| **Nadir CD4$^+$ count, cells/mm$^3$** | 229 (178) | 233 (178) | 222 (178) | 0.256 |
| **HIV viral load, copies/mL** | 20623 (75828) | 19744 (76284) | 22384 (74957) | 0.540 |
| **Soluble CD14 (sCD14), ng/mL** | 1810.3 (542.8) | 1804.3 (539.0) | 1822.4 (550.6) | 0.559 |
| **D-dimer, ng/mL** | 0.5 (1.0) | 0.5 (0.9) | 0.5 (1.1) | 0.536 |
| **Interleukin-6 (IL-6), pg/mL** | 3.2 (7.5) | 3.2 (8.0) | 3.4 (6.6) | 0.615 |
| **Primary outcome (incident CHF)** | 240 (17.2%) | 164 (17.6%) | 76 (16.3%) | 0.596 |

**Supplementary Table S2: Plasma abundance of proteins in final protein-only model.** †

| Protein name | Overall (N=1398) | Incident Heart Failure: No (N=1158) | Incident Heart Failure: Yes (N=240) | P-value |
|---|---|---|---|---|
| **Pancreatic ribonuclease** | 9.7 (0.9) | 9.6 (0.8) | 10.1 (1.3) | <0.001 |
| **WAP four-disulfide core domain protein 2** | 12.0 (0.6) | 11.9 (0.6) | 12.2 (0.7) | <0.001 |
| **Transgelin** | 12.5 (0.5) | 12.5 (0.4) | 12.7 (0.7) | <0.001 |
| **Fibulin 5** | 12.3 (0.3) | 12.3 (0.3) | 12.4 (0.3) | <0.001 |
| **Hepatitis A viral cellular receptor 1** | 9.8 (0.9) | 9.7 (0.8) | 10.2 (1.0) | <0.001 |
| **Adipocyte fatty acid-binding protein** | 14.2 (0.8) | 14.1 (0.7) | 14.5 (0.9) | <0.001 |
| **Macrophage metalloelastase** | 10.0 (0.7) | 10.0 (0.7) | 10.2 (0.6) | <0.001 |
| **Transmembrane glycoprotein NMB** | 9.9 (0.4) | 9.8 (0.4) | 10.0 (0.5) | <0.001 |

*Values presented as mean (SD) and p-values using the t-test*

| Protein name | Overall (N=1398) | Incident Heart Failure: No (N=1158) | Incident Heart Failure: Yes (N=240) | P-value |
|---|---|---|---|---|
| **Pancreatic ribonuclease** | 9.5 (9.1, 10.0) | 9.5 (9.1, 9.9) | 9.9 (9.3, 10.5) | <0.001 |
| **WAP four-disulfide core domain protein 2** | 11.9 (11.5, 12.3) | 11.9 (11.5, 12.3) | 12.2 (11.9, 12.6) | <0.001 |
| **Transgelin** | 12.5 (12.2, 12.8) | 12.5 (12.2, 12.7) | 12.6 (12.3, 13.0) | <0.001 |
| **Fibulin 5** | 12.3 (12.2, 12.5) | 12.3 (12.1, 12.4) | 12.4 (12.2, 12.6) | <0.001 |
| **Hepatitis A viral cellular receptor 1** | 9.6 (9.2, 10.2) | 9.6 (9.2, 10.1) | 10.0 (9.4, 10.7) | <0.001 |
| **Adipocyte fatty acid-binding protein** | 14.2 (13.7, 14.7) | 14.1 (13.6, 14.6) | 14.4 (13.9, 15.0) | <0.001 |
| **Macrophage metalloelastase** | 10.0 (9.5, 10.4) | 9.9 (9.5, 10.4) | 10.2 (9.8, 10.7) | <0.001 |
| **Transmembrane glycoprotein NMB** | 9.8 (9.6, 10.1) | 9.8 (9.6, 10.0) | 9.9 (9.7, 10.2) | <0.001 |

*Values presented as median (Q1, Q3) and p-values using the Mann Whitney U test*

† Protein values are reported in log2-normalized relative abundance units derived from SomaScan-normalized data after MAD-based outlier capping and log2 transformation. These are dimensionless normalized units (not absolute concentrations or relative fluorescence units); higher values indicate higher relative protein abundance.

| Supplementary Table S3: Baseline demographic and clinical characteristics of persons without HIV (PWoH) patients from VACS | |
|---|---|
| | PWoH |
| | (N=715) |
| Mean age, years (SD) | 53.8 (9.3) |
| Female, n (%) | 72 (10.1%) |
| Race/Ethnicity, n (%) | |
| White | 145 (20.3%) |
| Black | 475 (66.4%) |
| Hispanic | 62 (8.7%) |
| Other | 33 (4.6%) |
| Medical history, n (%) | |
| Hypertension | 488 (68.3%) |
| On antihypertensive treatment | 573 (80.1%) |
| Diabetes | 202 (28.3%) |
| Hyperlipidemia | 344 (48.1%) |
| On lipid-lowering therapy | 293 (41.0%) |
| Overweight/obese (BMI >25 kg/m$^2$) | 563 (79.0%) |
| Current Smoker | 346 (48.5%) |
| Cocaine use | 128 (19.1%) |
| Methamphetamine and other stimulant use | 14 (2.1%) |
| Prior ASCVD event | 50 (7.0%) |
| Mean (SD) systolic blood pressure, mmHg | 130.9 (13.9) |
| Mean (SD) body mass index, kg/m$^2$ | 29.9 (6.1) |
| On antiretroviral regimen at baseline, n (%) | -- |
| NRTI | -- |
| NNRTI | -- |
| Mean (SD) laboratory values) | |
| Total Cholesterol, mg/dL | 179.7 (44.6) |
| LDL Cholesterol, mg/dL | 104.8 (34.9) |
| HDL Cholesterol, mg/dL | 47.5 (15.1) |
| Estimated glomerular filtration rate, mL/min/1.73 m$^2$ | 97.0 (40.9) |
| CD4$^+$ count, cells/mm$^3$ | -- |
| Nadir CD4$^+$ count, cells/mm$^3$ | -- |
| HIV viral load, copies/mL | -- |
| Soluble CD14 (sCD14), ng/mL | -- |
| D-dimer, ng/mL | 0.5 (1.0) |
| Interleukin-6 (IL-6), pg/mL | 3.7 (15.7) |
| Primary outcome (incident heart failure event) | 143 (20.0%) |

**Supplementary Table S4: Predictive performance of multivariable models for incident heart failure among PWH and PWoH**

| Model | Covariates included in model | Proteins selected in model | Event-free survival C-index (95% confidence interval) | |
|---|---|---|---|---|
| | | | PWH test set, PWH-derived model (n = 466) | PWoH, PWH-derived model (n = 715) |
| | **CLINICAL RISK SCORES** | | | |
| 1 | ASCVD 10-year risk score[1] | -- | 0.607 (0.544 – 0.670) | 0.684 (0.641 – 0.727) |
| 2 | Clinical heart failure risk factors[2] | -- | 0.639 (0.574 – 0.704) | 0.707 (0.664 – 0.750) |
| 3a | Model 1 + eGFR[3] | -- | 0.607 (0.544 – 0.670) | 0.697 (0.654 – 0.740) |
| 3b | Model 2 + eGFR | -- | 0.640 (0.575 – 0.705) | 0.715 (0.674 – 0.756) |
| 4a | Model 1 + eGFR + stimulant use | -- | 0.630 (0.567 – 0.693) | 0.683 (0.640 – 0.726) |
| 4b | Model 2 + eGFR + stimulant use | -- | 0.647 (0.580 – 0.714) | 0.706 (0.663 – 0.749) |
| 5a | Model 3a + $CD4^+$ count + viral load | -- | 0.637 (0.576 – 0.698) | -- |
| 5b | Model 3b + $CD4^+$ count + viral load | -- | 0.656 (0.591 – 0.721) | -- |
| | **INFLAMMATORY MARKER RISK SCORES** | | | |
| 6 | sCD14 + IL-6 + D-dimer | -- | 0.611 (0.542 – 0.680) | 0.589 (0.542 – 0.636) |
| 7 | ASCVD score + sCD14 + IL-6 + D-dimer | -- | 0.632 (0.567 – 0.697) | 0.689 (0.646 – 0.732) |
| 8 | Clinical heart failure risk factors[2] + sCD14 + IL-6 + D-dimer | -- | 0.645 (0.580 – 0.710) | 0.718 (0.675 – 0.761) |
| 9 | Model 6 + $CD4^+$ count + viral load | -- | 0.600 (0.529 – 0.671) | -- |
| 10 | Model 7 + $CD4^+$ count + viral load | | 0.632 (0.567 – 0.697) | -- |
| 11 | Model 8 + $CD4^+$ count + viral load | | 0.657 (0.594 – 0.720) | -- |
| | **PROTEIN RISK SCORES** | | | |
| 12 | ASCVD score + **proteins** | RNASE1, NBL1, HE4, GAS1, FBLN3, TAGLN, FBLN5, TIM-1 | 0.713 (0.582 – 0.815) | 0.717 (0.625 – 0.794) |
| 13 | Clinical heart failure risk factors[2] + **proteins** | RNASE1, HE4, GAS1, TAGLN, FBLN5, TIM-1 | 0.717 (0.587 – 0.819) | 0.730 (0.639 – 0.806) |
| 14 | ASCVD score + sCD14 + D-dimer + IL-6 + **proteins** | RNASE1, NBL1, HE4, GAS1, TAGLN, FBLN5, TIM-1 | 0.710 (0.579 – 0.814) | 0.716 (0.624 – 0.794) |
| 15 | **Proteins only** | RNASE1, HE4, TAGLN, FBLN5, TIM-1, FABPA, MMP-12, GPNMB | 0.725 (0.597 – 0.825) | 0.699 (0.608 – 0.777) |
| 16 | **Proteins** + $CD4^+$ count + viral load | RNASE1, HE4, PXDN, TAGLN, FBLN5, TIM-1, GPNMB | 0.715 (0.587 – 0.816) | -- |
| 17 | **Proteins** + $CD4^+$ count + viral load + sCD14 + IL-6 + D-dimer | RNASE1, HE4, PXDN, TAGLN, FBLN5, TIM-1, GPNMB | 0.720 (0.591 – 0.820) | -- |

[1] ACC/AHA Pooled Cohort Equations

[2] Age; sex; race; systolic blood pressure; antihypertensive medication use; diabetes; current smoker; body mass index; total cholesterol; high-density lipoprotein cholesterol

[3] eGFR, estimated glomerular filtration rate

[4] sCD14, soluble CD14; IL-6, interleukin-6

C-index

0.60 0.65 0.70

**Supplementary Table S5: Candidate protein markers (m = 108) validated in test set in order of descending significance (Bonferroni-corrected threshold: p <1E-5). Color scale represents magnitude and direction of hazard ratio (HR) for incident heart failure. The 8 proteins selected in the final protein model are highlighted in magenta. Druggable targets are proteins whose structures have been characterized.**

| UniProt ID | Protein | Gene(s) | Druggable target | Hazard ratio (95% confidence interval) |
|---|---|---|---|---|
| **P07998** | **Ribonuclease pancreatic** | **RNASE1 RIB1 RNS1** | | **1.75 (1.48 - 2.08)** |
| P41271 | Neuroblastoma suppressor of tumorigenicity 1 | NBL1 DAN DAND1 | | 2.06 (1.67 - 2.52) |
| **Q14508** | **WAP four-disulfide core domain protein 2** | **WFDC2 HE4 WAP5** | | **2.92 (2.03 - 4.20)** |
| Q92626 | Peroxidasin homolog | PXDN KIAA0230 MG50 PRG2 PXD01 VPO VPO1 | | 1.88 (1.56 - 2.28) |
| O95633 | Follistatin-related protein 3 | FSTL3 FLRG UNQ674/PRO1308 | Yes | 3.21 (2.14 - 4.81) |
| P01034 | Cystatin-C | CST3 | Yes | 3.28 (2.26 - 4.77) |
| P19438 | Tumor necrosis factor receptor superfamily member 1A | TNFRSF1A TNFAR TNFR1 | Yes | 2.99 (2.17 - 4.11) |
| Q07654 | Trefoil factor 3 | TFF3 ITF TFI | | 1.83 (1.42 - 2.36) |
| P54826 | Growth arrest-specific protein 1 | GAS1 | | 3.15 (1.95 - 5.08) |
| Q12805 | EGF-containing fibulin-like extracellular matrix protein 1 | EFEMP1 FBLN3 FBNL | | 3.31 (2.24 - 4.88) |
| **Q01995** | **Transgelin** | **TAGLN SM22 WS3-10** | **Yes** | **2.44 (1.71 - 3.48)** |
| P49755 | Transmembrane emp24 domain-containing protein 10 | TMED10 TMP21 | | 1.95 (1.41 - 2.71) |
| P17900 | Ganglioside GM2 activator | GM2A | Yes | 2.73 (1.97 - 3.78) |
| P12111 | Collagen alpha-3(VI) chain | COL6A3 | Yes | 2.71 (1.95 - 3.77) |
| O43921 | Ephrin-A2 | EFNA2 EPLG6 LERK6 | Yes | 2.53 (1.68 - 3.81) |
| P52798 | Ephrin-A4 | EFNA4 EPLG4 LERK4 | Yes | 2.63 (1.84 - 3.77) |
| Q2UY09 | Collagen alpha-1(XXVIII) chain | COL28A1 COL28 | | 3.04 (2.14 - 4.31) |
| P61769 | Beta-2-microglobulin | B2M CDABP0092 HDCMA22P | Yes | 2.25 (1.67 - 3.03) |
| Q01974 | Tyrosine-protein kinase transmembrane receptor ROR2 | ROR2 NTRKR2 | Yes | 2.46 (1.85 - 3.29) |
| **Q9UBX5** | **Fibulin-5** | **FBLN5 DANCE UNQ184/PRO210** | | **3.65 (1.85 - 7.19)** |
| P29317 | Ephrin type-A receptor 2 | EPHA2 ECK | Yes | 2.77 (2.05 - 3.73) |
| Q8IZJ1 | Netrin receptor UNC5B | UNC5B P53RDL1 UNC5H2 UNQ1883/PRO4326 | | 3.29 (2.10 - 5.15) |
| Q02487 | Desmocollin-2 | DSC2 CDHF2 DSC3 | | 2.24 (1.59 - 3.16) |
| P52803 | Ephrin-A5 | EFNA5 EPLG7 LERK7 | | 2.91 (1.87 - 4.52) |
| Q8IZJ1 | Netrin receptor UNC5B | UNC5B P53RDL1 UNC5H2 UNQ1883/PRO4326 | | 3.67 (2.36 - 5.70) |
| Q8TDQ0 | Hepatitis A virus cellular receptor 2 | HAVCR2 TIM3 TIMD3 | Yes | 3.27 (2.26 - 4.75) |
| P20333 | Tumor necrosis factor receptor superfamily member 1B | TNFRSF1B TNFBR TNFR2 | Yes | 2.77 (1.86 - 4.11) |
| Q6UY11 | Protein delta homolog 2 | DLK2 EGFL9 UNQ2903/PRO28633 | | 2.86 (2.00 - 4.10) |

HR

0.25 1.0 5.0

**Supplementary Table S5: Candidate protein markers (m = 108) validated in test set in order of descending significance (Bonferroni-corrected threshold: p <1E-5). Color scale represents magnitude and direction of hazard ratio (HR) for incident heart failure. The 8 proteins selected in the final protein model are highlighted in magenta. Druggable targets are proteins whose structures have been characterized.**

| UniProt ID | Protein | Gene(s) | Druggable target | Hazard ratio (95% confidence interval) |
|---|---|---|---|---|
| Q13261 | Interleukin-15 receptor subunit alpha | IL15RA | Yes | 2.32 (1.63 - 3.30) |
| P21246 | Pleiotrophin | PTN HBNF1 NEGF1 | Yes | 3.31 (1.96 - 5.58) |
| P20333 | Tumor necrosis factor receptor superfamily member 1B | TNFRSF1B TNFBR TNFR2 | | 2.32 (1.62 - 3.33) |
| P52799 | Ephrin-B2 | EFNB2 EPLG5 HTKL LERK5 | | 2.68 (1.77 - 4.06) |
| Q8N2S1 | Latent-transforming growth factor beta-binding protein 4 | LTBP4 | | 2.59 (1.64 - 4.11) |
| Q8WWX9 | Selenoprotein M | SELENOM SELM | | 3.28 (2.15 - 5.01) |
| Q9NP99 | Triggering receptor expressed on myeloid cells 1 | TREM1 | Yes | 1.93 (1.48 - 2.52) |
| Q9BUD6 | Spondin-2 | SPON2 DIL1 UNQ435/PRO866 | | 2.71 (1.82 - 4.02) |
| Q7Z4F1 | Low-density lipoprotein receptor-related protein 10 | LRP10 MSTP087 SP220 UNQ389/PRO724 | | 4.16 (2.61 - 6.65) |
| Q9GZN4 | Brain-specific serine protease 4 | PRSS22 BSSP4 PRSS26 SP001LA UNQ302/PRO343 | | 2.14 (1.46 - 3.13) |
| Q8N6G6 | ADAMTS-like protein 1 | ADAMTSL1 ADAMTSR1 C9orf94 UNQ528/PRO1071 | | 3.73 (2.19 - 6.36) |
| Q93091 | Ribonuclease K6 | RNASE6 RNS6 | | 2.32 (1.55 - 3.47) |
| O15123 | Angiopoietin-2 | ANGPT2 | Yes | 2.80 (1.98 - 3.96) |
| Q2TAL6 | Brorin | VWC2 UNQ739/PRO1434 | | 2.69 (1.61 - 4.52) |
| **Q96D42** | **Hepatitis A virus cellular receptor 1** | **HAVCR1 KIM1 TIM1 TIMD1** | | **1.76 (1.48 - 2.11)** |
| P00533 | Epidermal growth factor receptor | EGFR ERBB ERBB1 HER1 | Yes | 0.21 (0.09 - 0.46) |
| O00478 | Butyrophilin subfamily 3 member A3 | BTN3A3 BTF3 | | 1.48 (1.20 - 1.82) |
| P07306 | Asialoglycoprotein receptor 1 | ASGR1 CLEC4H1 | Yes | 2.51 (1.59 - 3.97) |
| Q86VZ4 | Low-density lipoprotein receptor-related protein 11 | LRP11 | | 3.54 (1.89 - 6.63) |
| Q9UBX7 | Kallikrein-11 | KLK11 PRSS20 TLSP UNQ649/PRO1279 | | 2.78 (1.73 - 4.47) |
| Q9H665 | IGF-like family receptor 1 | IGFLR1 TMEM149 U2AF1L4 | | 1.81 (1.48 - 2.20) |
| O00292 | Left-right determination factor 2 | LEFTY2 EBAF LEFTA LEFTYA TGFB4 PSEC0024 | | 2.88 (1.74 - 4.76) |
| P16860 | N-terminal pro-BNP | NPPB | Yes | 1.94 (1.62 - 2.32) |
| P13987 | CD59 glycoprotein | CD59 MIC11 MIN1 MIN2 MIN3 MSK21 | Yes | 2.52 (1.63 - 3.90) |
| O43155 | Leucine-rich repeat transmembrane protein FLRT2 | FLRT2 KIAA0405 UNQ232/PRO265 | | 3.36 (1.92 - 5.88) |
| Q03403 | Trefoil factor 2 | TFF2 SML1 | | 2.07 (1.53 - 2.80) |
| Q9H3T3 | Semaphorin-6B | SEMA6B SEMAN SEMAZ UNQ1907/PRO4353 | | 2.69 (1.85 - 3.91) |
| P54760 | Ephrin type-B receptor 4 | EPHB4 HTK MYK1 TYRO11 | Yes | 2.91 (1.81 - 4.68) |
| Q01105 | Protein SET | SET | | 0.19 (0.08 - 0.44) |
| O14558 | Heat shock protein beta-6 | HSPB6 | | 2.35 (1.65 - 3.33) |

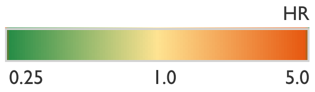

**Supplementary Table S5: Candidate protein markers (m = 108) validated in test set in order of descending significance (Bonferroni-corrected threshold: p <1E-5). Color scale represents magnitude and direction of hazard ratio (HR) for incident heart failure. The 8 proteins selected in the final protein model are highlighted in magenta. Druggable targets are proteins whose structures have been characterized.**

| UniProt ID | Protein | Gene(s) | Druggable target | Hazard ratio (95% confidence interval) |
|---|---|---|---|---|
| **P15090** | **Fatty acid-binding protein, adipocyte** | **FABP4** | **Yes** | **2.13 (1.58 - 2.86)** |
| Q9UGN4 | CMRF35-like molecule 8 | CD300A CMRF35H IGSF12 HSPC083 | Yes | 2.23 (1.55 - 3.20) |
| P21757 | Macrophage scavenger receptor types I and II | MSR1 SCARA1 | Yes | 2.12 (1.64 - 2.73) |
| Q96DX5 | Ankyrin repeat and SOCS box protein 9 | ASB9 | | 2.68 (1.71 - 4.21) |
| O95150 | Tumor necrosis factor ligand superfamily member 15 | TNFSF15 TL1 VEGI | Yes | 1.82 (1.43 - 2.31) |
| P15529 | Membrane cofactor protein | CD46 MCP MIC10 | Yes | 4.80 (2.51 - 9.19) |
| Q8IYJ0 | PILR alpha-associated neural protein | PIANP C12orf53 PANP UNQ828/PRO1755 | | 1.65 (1.27 - 2.15) |
| P35442 | Thrombospondin-2 | THBS2 TSP2 | | 1.49 (1.22 - 1.83) |
| O43915 | Vascular endothelial growth factor D | VEGFD FIGF | Yes | 2.08 (1.41 - 3.06) |
| Q6PUV4 | Complexin-2 | CPLX2 | | 1.95 (1.44 - 2.63) |
| A6NI73 | Leukocyte immunoglobulin-like receptor subfamily A member 5 | LILRA5 ILT11 LILRB7 LIR9 | Yes | 2.93 (1.69 - 5.10) |
| P84157 | Matrix-remodeling-associated protein 7 | MXRA7 | | 1.80 (1.38 - 2.35) |
| Q9BT09 | Protein canopy homolog 3 | CNPY3 CTG4A ERDA5 PRAT4A TNRC5 HSPC084 UNQ1934/PRO4409 | | 2.52 (1.71 - 3.73) |
| P08949 | Neuromedin-B | NMB | | 3.11 (1.88 - 5.16) |
| Q13753 | Laminin subunit gamma-2 | LAMC2 LAMB2T LAMNB2 | Yes | 2.19 (1.55 - 3.09) |
| P17174 | Aspartate aminotransferase, cytoplasmic | GOT1 | Yes | 0.15 (0.07 - 0.36) |
| Q08708 | CMRF35-like molecule 6 | CD300C CMRF35 CMRF35A CMRF35A1 IGSF16 | | 2.34 (1.67 - 3.28) |
| O60259 | Kallikrein-8 | KLK8 NRPN PRSS19 TADG14 UNQ283/PRO322 | Yes | 2.91 (1.76 - 4.81) |
| Q86VZ4 | Low-density lipoprotein receptor-related protein 11 | LRP11 | | 3.21 (2.01 - 5.13) |
| Q9UHX3 | Adhesion G protein-coupled receptor E2 | ADGRE2 EMR2 | | 2.25 (1.60 - 3.16) |
| O00300 | Tumor necrosis factor receptor superfamily member 11B | TNFRSF11B OCIF OPG | Yes | 3.09 (2.09 - 4.58) |
| P05451 | Lithostathine-1-alpha | REG1A PSPS PSPS1 REG | | 1.68 (1.29 - 2.19) |
| Q9Y4C0 | Neurexin-3 | NRXN3 C14orf60 KIAA0743 | | 1.94 (1.49 - 2.53) |
| Q16832 | Discoidin domain-containing receptor 2 | DDR2 NTRKR3 TKT TYRO10 | Yes | 1.60 (1.26 - 2.04) |
| P58340 | Myeloid leukemia factor 1 | MLF1 | | 0.13 (0.05 - 0.33) |
| Q9H8J5 | MANSC domain-containing protein 1 | MANSC1 LOH12CR3 UNQ316/PRO361 | | 2.68 (1.70 - 4.23) |
| Q86TH1 | ADAMTS-like protein 2 | ADAMTSL2 KIAA0605 | | 1.80 (1.37 - 2.37) |
| O75509 | Tumor necrosis factor receptor superfamily member 21 | TNFRSF21 DR6 UNQ437/PRO868 | Yes | 3.89 (2.32 - 6.54) |

HR

0.25 1.0 5.0

**Supplementary Table S5: Candidate protein markers (m = 108) validated in test set in order of descending significance (Bonferroni-corrected threshold: p <1E-5). Color scale represents magnitude and direction of hazard ratio (HR) for incident heart failure. The 8 proteins selected in the final protein model are highlighted in magenta. Druggable targets are proteins whose structures have been characterized.**

| UniProt ID | Protein | Gene(s) | Druggable target | Hazard ratio (95% confidence interval) |
|---|---|---|---|---|
| Q9UJJ9 | N-acetylglucosamine-1-phosphotransferase subunit gamma | GNPTG C16orf27 GNPTAG CAB56184 LP2537 | | 3.39 (1.80 - 6.38) |
| Q9BRR6 | ADP-dependent glucokinase | ADPGK PSEC0260 | | 4.25 (2.15 - 8.39) |
| O00244 | Copper transport protein ATOX1 | ATOX1 HAH1 | | 1.58 (1.25 - 2.00) |
| **P39900** | **Macrophage metalloelastase 12** | **MMP12 HME** | **Yes** | **1.90 (1.45 - 2.49)** |
| P04070 | Vitamin K-dependent protein C | PROC | Yes | 0.28 (0.17 - 0.47) |
| Q01973 | Inactive tyrosine-protein kinase transmembrane receptor ROR1 | ROR1 NTRKR1 | Yes | 2.03 (1.42 - 2.90) |
| Q13291 | Signaling lymphocytic activation molecule | SLAMF1 SLAM | Yes | 1.77 (1.33 - 2.36) |
| P30041 | Peroxiredoxin-6 | PRDX6 AOP2 KIAA0106 | | 0.33 (0.19 - 0.58) |
| **Q14956** | **Transmembrane glycoprotein NMB** | **GPNMB HGFIN NMB UNQ1725/PRO9925** | **Yes** | **2.90 (1.96 - 4.29)** |
| Q9NZC2 | Triggering receptor expressed on myeloid cells 2 | TREM2 | Yes | 2.00 (1.50 - 2.66) |
| Q9HDB5 | Neurexin-3-beta | NRXN3 KIAA0743 | | 1.94 (1.50 - 2.51) |
| P22223 | Cadherin-3 | CDH3 CDHP | Yes | 0.35 (0.20 - 0.59) |
| Q9GZX9 | Twisted gastrulation protein homolog 1 | TWSG1 TSG PSEC0250 | | 4.03 (2.10 - 7.75) |
| Q6IE38 | Serine protease inhibitor Kazal-type 14 | SPINK14 SPINK5L2 | | 2.41 (1.52 - 3.81) |
| Q9NR61 | Delta-like protein 4 | DLL4 UNQ1895/PRO4341 | Yes | 3.35 (2.07 - 5.42) |
| Q9NS68 | Tumor necrosis factor receptor superfamily member 19 | TNFRSF19 TAJ TROY UNQ1888/PRO4333 | | 2.06 (1.49 - 2.85) |
| P01024 | C3a anaphylatoxin | C3 CPAMD1 | | 0.19 (0.09 - 0.42) |
| Q8N3J6 | Cell adhesion molecule 2 | CADM2 IGSF4D NECL3 | | 2.33 (1.55 - 3.50) |
| P31415 | Calsequestrin-1 | CASQ1 CASQ | | 0.15 (0.06 - 0.37) |
| Q8NBI6 | Xyloside xylosyltransferase 1 | XXYLT1 C3orf21 PSEC0251 | | 5.10 (2.72 - 9.54) |
| Q6EMK4 | Vasorin | VASN SLITL2 UNQ314/PRO357/PRO1282 | | 4.01 (2.13 - 7.55) |
| P17936 | Insulin-like growth factor-binding protein 3 | IGFBP3 IBP3 | Yes | 0.36 (0.24 - 0.54) |

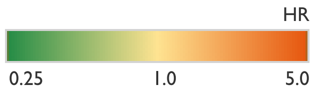

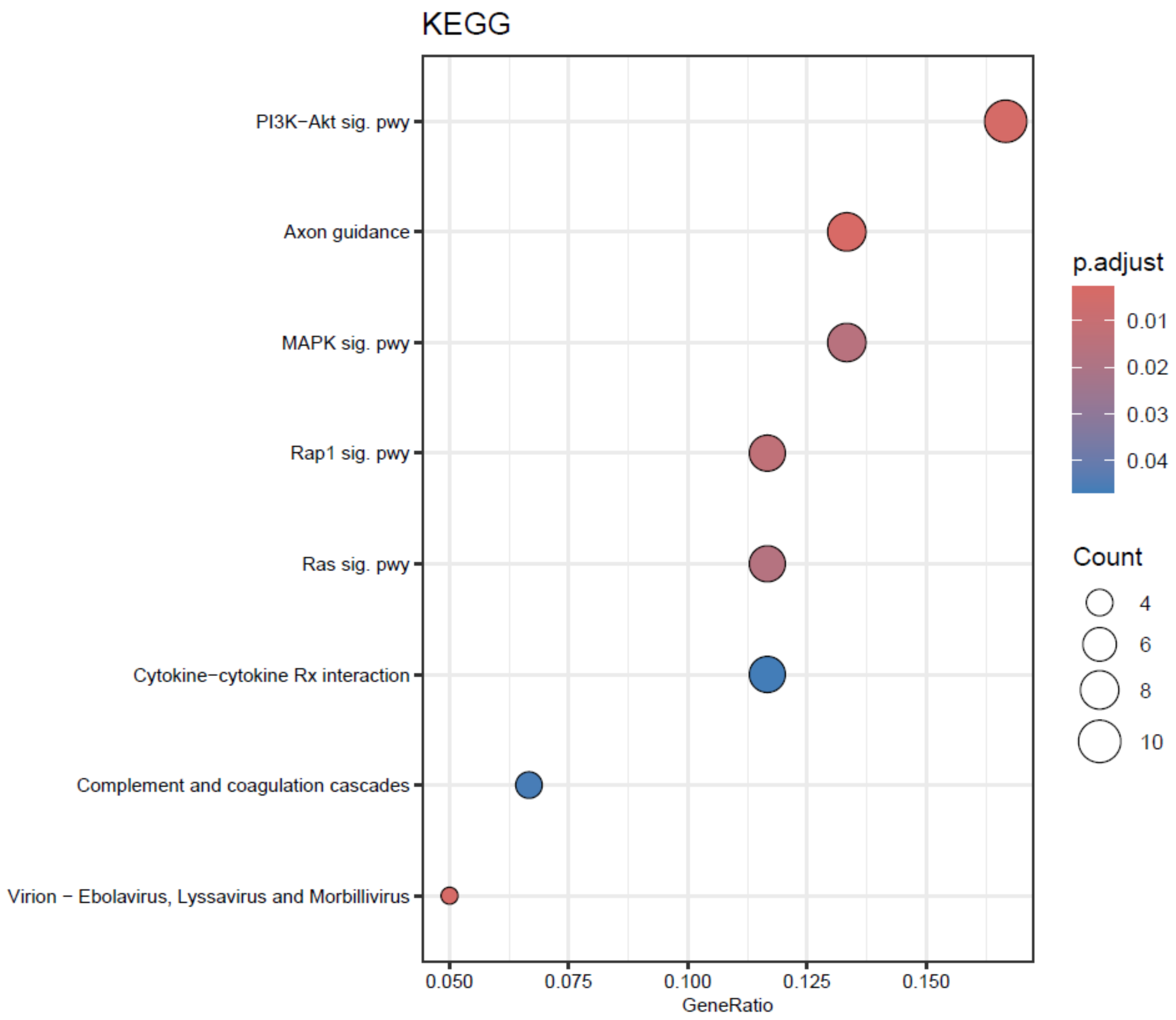


**Supplementary Figure S1: Results of Kyoto Encyclopedia of Genes and Genomes (KEGG) pathway enrichment analysis for 108 candidate proteins for final model.**

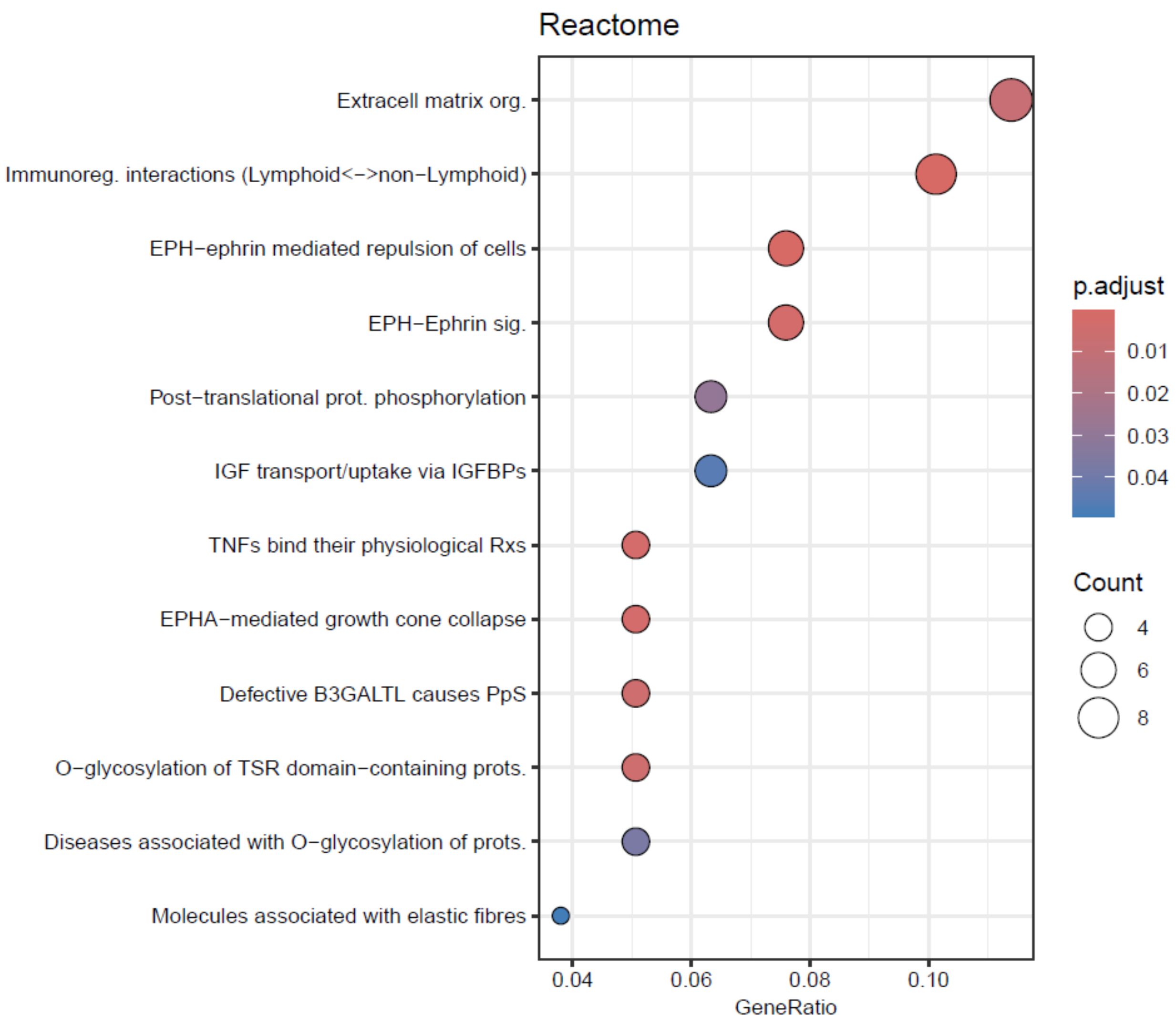


**Supplementary Figure S2: Results of Reactome pathway enrichment analysis for 108 candidate proteins for final model.**

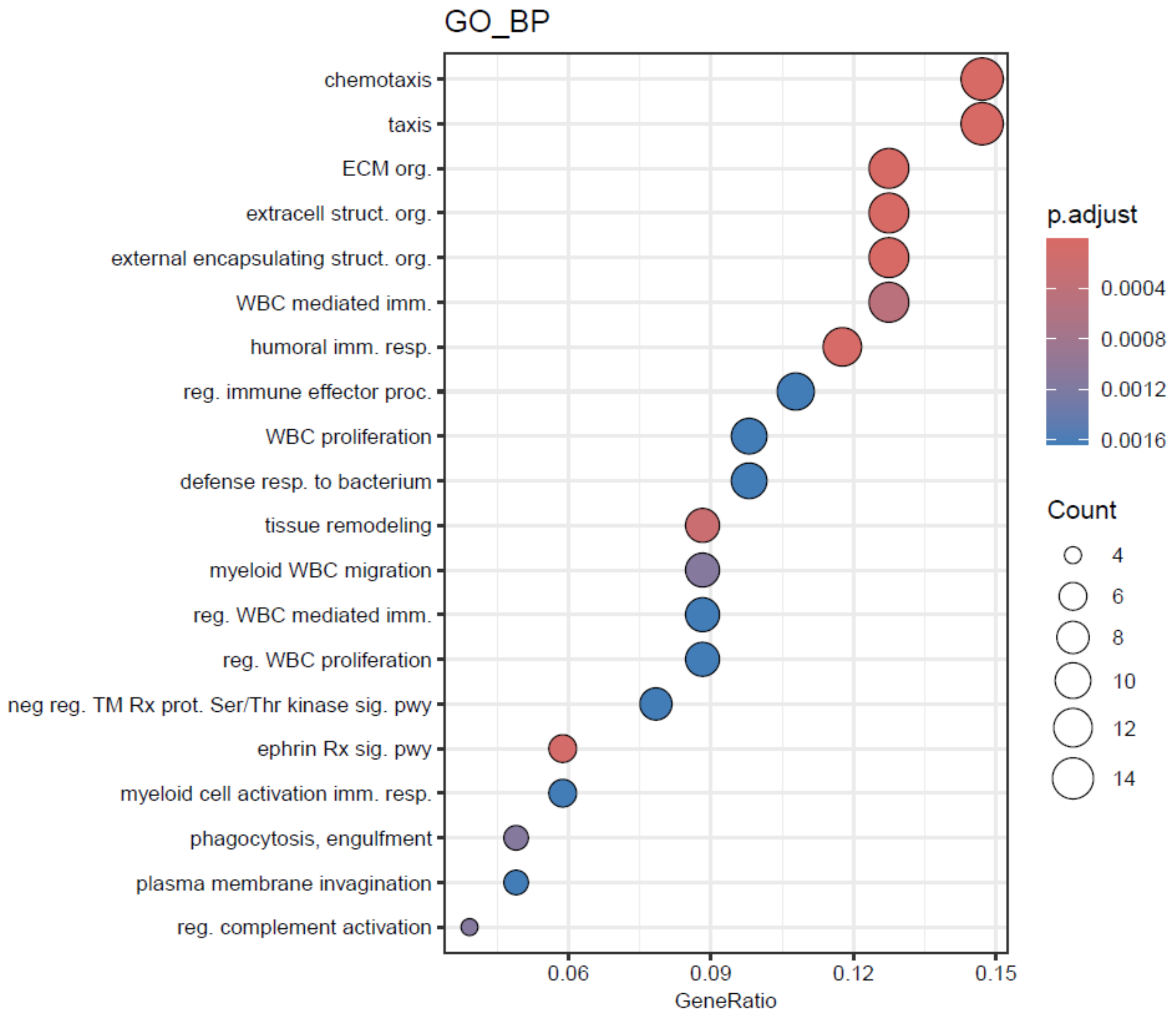


**Supplementary Figure S3: Results of Gene Ontology Biological Process (GO BP) pathway enrichment analysis for 108 candidate proteins for final model.**
